# Prevalence of COVID-19 in Iran: Results of the first survey of the Iranian COVID-19 Serological Surveillance program

**DOI:** 10.1101/2021.03.12.21253442

**Authors:** Kazem Khalagi, Safoora Gharibzadeh, Davood Khalili, Mohammad Ali Mansournia, Siamak Mirab Samiee, Saeide Aghamohamadi, Maryam Mir-Mohammad-Ali Roodaki, Seyed Mahmoud Hashemi, Katayoun Tayeri, Hengameh Namdari Tabar, Kayhan Azadmanesh, Jafar Sadegh Tabrizi, Kazem Mohammad, Firoozeh Hajipour, Saeid Namaki, Alireza Raeisi, Afshin Ostovar

## Abstract

**Background:** This study aims to estimate the prevalence of COVID-19 in the general population of Iran.

**Methods:** The target population was all Iranian people aged six years and older in the country. A stratified random sampling design was used to select 28,314 subjects from among the individuals registered in the electronic health record systems used in primary health care in Iran. Venous blood was taken from each participant and tested for the IgG antibody against COVID-19. The prevalence of COVID-19 was estimated at provincial and national levels after adjusting for the measurement error of the laboratory test, non-response bias, and sampling design.

**Results:** Of the 28,314 Iranians selected, 11,256 (39.75%) participated in the study. Of these, 5406 (48.0%) were male, and 6851 (60.9%) lived in urban areas. The mean (standard deviation) participant age was 35.89 (18.61) years. The adjusted prevalence of COVID-19 until August 20, 2020 was estimated as 14.2% (95% uncertainty interval: 13.3%, 15.2%), which was equal to 11,958,346 (95% confidence interval: 11,211,011–12,746,776) individuals. The prevalence of infection was 14.6%, 13.8%, 16.6%, 11.7%, and 19.4% among men, women, urban population, rural population, and individuals ≥60 years of age, respectively. Ardabil, Golestan, and Khuzestan provinces had the highest prevalence, and Alborz, Hormozgan, and Kerman provinces had the lowest.

**Conclusions:** Based on the study results, a large proportion of the Iranian population had not yet been infected by COVID-19. The observance of hygienic principles and social restrictions should therefore continue until the majority of the population has been vaccinated.

## Introduction

The severe acute respiratory syndrome coronavirus 2 (SARS-CoV-2) pandemic, which started in December 2019 in Wuhan, China, and quickly spread around the world, is one of the most important phenomena that has affected human society in recent decades (1). The agent of SARS-CoV-2 was a RNA beta-coronavirus that had never been seen before (2). Due to the rapid transmission of infection from person to person via respiration, measures such as social distancing and lockdowns have been taken in different communities and countries to control the pandemic. These have not only affected people’s physical health, but also their social and mental health and have had a significantly negative effect on the economies of families and countries. Accurate and valid information about the prevalence of infection (symptomatic and asymptomatic) is important for policy-making and the management and control of the COVID-19 pandemic in countries (3). Although daily reports of the number of polymerase chain reaction (PCR)-confirmed cases of COVID-19 are available, the number of diagnosed cases is a function of the number of tests performed per day. For example, in Iran, due to the limited number of laboratory kits available at the beginning of the pandemic, the number of daily tests was very limited, and PCR tests were performed only for hospitalized patients in a serious physical condition with suspected COVID-19 (4). Furthermore, the PCR test has a relatively high false-negative rate, which is affected by the sampling method and the time interval from the onset of the disease (5). On the other hand, a significant proportion (17%) of COVID-19 patients remain asymptomatic, especially those in younger age groups (6, 7). Given the above limitations, it is impossible to rely solely on daily reports of the number of definitively diagnosed cases for pandemic management and policy-making.

Serological tests are used to measure the response of antibodies to the virus and are also able to detect a history of infection in asymptomatic individuals. Seroepidemiological surveys can therefore provide reliable information about the prevalence of the infection, its distribution in different areas and by age, sex, and other subgroups, and the history of population immunity (8-10).

To date, several seroprevalence studies have been performed in metropolises and provinces in Iran and on high-risk populations, and some of these results have been published (11, 12). However, the surveys have had a number of limitations in terms of the sampling designs and analysis methods, which has raised debate on the validity and generalizability of the findings (13-15). This study from the Iranian COVID-19 Serological Surveillance (ICS) program, which is supported by the Ministry of Health and Medical Education (MOHME) of Iran (16), is the first report of the series of nationwide, population-based serological surveys for COVID-19 that are conducted at regular intervals.

## Methods

### Study design, population, and sampling

The survey was conducted from August to October 2020 in all provinces of Iran to estimate the prevalence of COVID-19 in the country in total and by province, urban/rural area of residence, sex, and age group. The target population of the survey was all Iranians aged six years and older living in Iran. People with a unique Iranian national identification number registered in the primary health care (PHC) electronic health record systems (SIB, SINA, and NAB), who were six years of age or older and had sufficient physical ability to attend blood sampling centers were included. Subjects who had contraindications for venous blood sampling or were not willing to participate in the study were excluded.

A stratified random sampling scheme was used in the survey; each province was considered a stratum. In each province, sampling was conducted through a simple random sampling method using the list of eligible subjects registered in the PHC electronic health record systems as the sampling frame.

A national sample of 28,314 individuals was recruited. The sample included 858 subjects for each of the 31 provinces in Iran with the exception of Tehran province. For Tehran, a three times greater number of participants were recruited compared to the sample size calculated for each of the other provinces. The provincial sample size was calculated based on the estimated COVID-19 prevalence of 33% (17), a relative estimation error of 10%, a confidence interval (CI) of 95%, and considering a non-response rate of 10%. The country sample size was sufficient to estimate a prevalence of 33% with an estimation error of 1.75%.

### Procedures

To invite the subjects selected for the survey, the relevant lists and personal profiles in the PHC electronic health record systems were made available to urban and rural community health workers. These community health workers called the selected people and invited them to each district’s specified blood sampling centers for blood sampling. If the community health workers failed to reach the subjects by phone, this would be repeated up to three times and up to twice a day. Individuals who gave verbal consent to participate in the study were asked to visit the relevant blood sampling center of the district within a maximum of five working days. If they did not attend the sampling center, the community health worker was informed through the system and contacted them again to follow up.

The blood sampling centers in each district were selected in such a way that the participants would face minimal risk of being infected by COVID-19 and the centers were easily accessible to the participants. Blood sampling was performed in full compliance with the health protocols and after obtaining written informed consent from the participants. A volume of 10 ml of intravenous blood was taken from each person, and the unique code of the person in the PHC electronic health record system, the names of the district and province, and the date of sampling were recorded on the sample tubes. Up to two hours after blood sampling, the sample tubes were centrifuged at 1000–1200 rpm for a maximum of 15 minutes, and the serum was then transferred to plastic-sealed micro tubes, which were stored at 4°C–8°C. The serum samples were then transferred to the selected laboratories of the medical universities in a three-layer package at a temperature of 4°C–8°C up to 24 hours after sampling.

### Measurements and other variables

To determine the IgG antibody against COVID-19 of each sample, serological testing was undertaken via the ELISA method using Iran’s Food and Drug Organization-approved SARS-CoV-2 ELISA kit (Pishtaz Teb, Tehran, Iran; catalogue number PT-SARS-COV-2.IgG-96) according to the relevant protocol. The test result was then recorded by the laboratory staff in the PHC electronic health record system. Other required variables such as age, gender, province/district of residence, and urban/rural area of residence were extracted from the participant’s profile in the PHC electronic health record system. Figure S1 of Appendix A shows the study implementation process.

### Ethical considerations

The study protocol was approved by the ethics committee of the National Institute of Health Research of the Islamic Republic of Iran (ethics code: IR.TUMS.NIHR.REC.1399.019). Written informed consent was obtained from all the participants in the study. For the participants aged 12–18 years, in addition to their individual consent, the consent of their parents or legal guardians was obtained. For children under 12 years of age, written informed consent was obtained only from their parents or legal guardians.

### Education, monitoring, and supervision

The objectives and protocols of the survey were described to the focal points of the health laboratories at the medical universities/faculties via a virtual training session, and they were asked to transfer the training materials hierarchically to community health workers, blood samplers, and laboratory personnel.

All the study processes were monitored using two tools: (i) the dashboard of the PHC electronic health record systems to assess the progress of the study and (ii) on-site checklists. The directors of the health laboratories of the medical universities were responsible for supervising the study processes.

### Estimations of the laboratory kit sensitivity and specificity

In addition to the reports of the diagnostic accuracy of the Pishtaz Teb kit by the manufacturer, its accuracy was reevaluated in a separate study. To assess the sensitivity of the kit, 254 patients with PCR-confirmed diagnoses of COVID-19 were tested (18). These individuals comprised patients admitted to hospital, those attending outpatient clinics, and asymptomatic subjects (19). Symptomatic patients were selected from among those whose symptoms had appeared at least three weeks earlier.

To estimate the specificity of the kit, we used 410 healthy people’s serum samples from the bio-bank of the Tehran Lipid and Glucose Study (20), which had been stored one year prior to the onset of the COVID-19 pandemic (fall 2018 to summer 2019). The serum samples consisted of a combination of samples taken during all the seasons of the year in equal proportions. We used multiple modified Poisson regression to evaluate the factors affecting the sensitivity and specificity of the study kit (21).

### Statistical analysis

We estimated the prevalence of COVID-19 and its 95% uncertainty interval (UI) at a national and provincial level by urban/rural area of residence, sex, and age (6–17 years, 18–39 years, 40– 59 years, and ≥60 years). A participant was considered “positive” in the presence of anti-SARS-CoV-2 IgG. After correcting the false negatives and false positives of the IgG test results and using post-stratification, inverse probability of response, and sampling design weights, a minimum bias estimate of the prevalence was obtained.

We followed three stages in the statistical analysis: (i) correction of the crude (unadjusted) prevalence resulting from the measurement error of the laboratory kit based on the sensitivity and specificity of the kit, (ii) conversion of the corrected prevalence of the previous stage into individual data, (iii) weighing the individual data of the second stage using post-stratification, response rates, and sampling design weights. All these stages were performed for 16 strata made up of a combination of four age groups, two genders, and two urban/rural categories in each province separately (496 categories at a national level).

To correct the crude prevalence resulting from the measurement error of the laboratory kit, we used the Bayesian method. In this method, the beta distributions of sensitivity and specificity and the uniform (0 and 1) distribution of the crude prevalence were used as three prior distributions, and the posterior distribution of the prevalence was then obtained using equation (1):

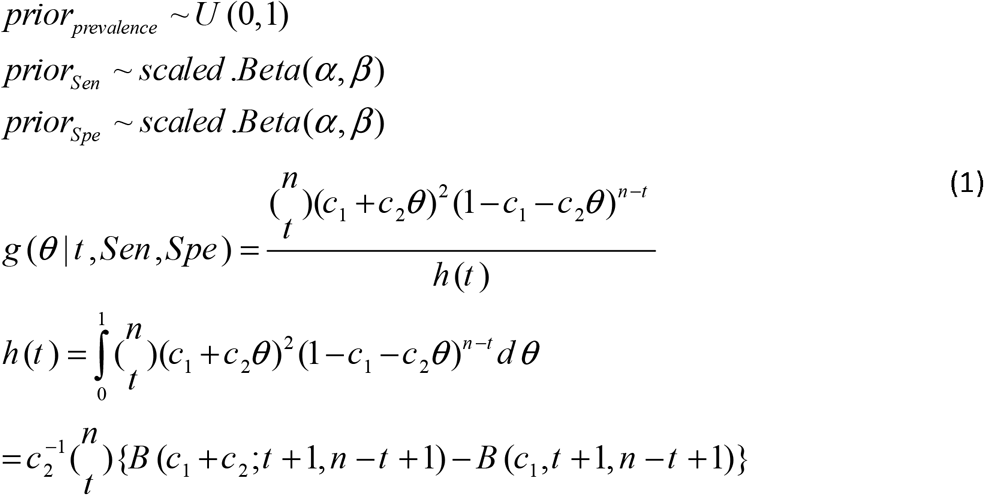

where □ is the crude prevalence, *t* is the number of positive subjects from *n* people tested, *C*_*1*_*=1-Spe*, and *C*_*2*_*=Sen + Spe −1* (22).

The beta distributions of the sensitivity and specificity were constructed in such a way that their means and standard deviations could be matched with the point estimates and standard errors of the sensitivity and specificity obtained from the kit’s performance study, respectively.

In stage 2, to convert the corrected prevalence of the previous stage into individual data, we simulated the individual data using a binomial distribution with parameters equal to the number of participants and the corrected prevalence of each of the aforementioned 16 subgroups in each province.

In stage 3, to obtain the prevalence estimates at a provincial level, we used the following two weights:

1. The weight of the corrected differences in the age, sex, and urban/rural distribution of the study sample with their distribution in the population of the province (W1 or post-stratification weight). This weight was the inverse of the ratio of the number of samples determined for each age-sex-urban/rural category by the population of that category in each province based on the population projection for 2020 by the Statistics Center of Iran;
2. The weight of the responses (W2). This weight was used to correct the effect of non-responses on the prevalence estimates (23). Since the variables age, sex, urban/rural area, and province of residence were associated with participation in the study, W2 was obtained by dividing the number of determined samples by the number of participants (inverse of probability of responses) in each of the 16 age-sex-urban/rural categories for each province.

We used the product of W1 and W2 as a weight to calculate the prevalence estimates in each province.

To obtain the prevalence estimates at a national level, W3 was calculated by dividing the number of the population of each province by the sample size of that province (sampling weight) (23). To calculate the national prevalence estimates, W3 was multiplied by W1 and W2. The standard error of the corrected prevalence was calculated using the robust method (24). Microsoft Excel (Microsoft Inc), STATA (25) and R (26) software were used in the statistical analysis.

The full details of the survey methods are also provided in an ICS program protocol article, which has been published elsewhere (16).

## Results

Among the 28,314 Iranians selected to participate in the study from all the provinces in the country, 11,256 (39.75%) participated in the study (Figure 1). The highest rate of non-response was reported from Tehran, Qom, and Hamadan provinces (90.4%, 76.9%, and 76.5%, respectively), while the provinces of Razavi Khorasan, Sistan and Baluchestan, and Mazandaran had the lowest non-response rates (14.7%, 24.2%, and 33.7%, respectively; see Table 1). The multiple logistic regression analysis used to investigate the factors affecting participation in the study revealed a statistically significant association between study participation and province of residence (odds ratio [OR]: 1.074, 95% CI: 1.071–1.077), one-year aging (OR: 1.007, 95% CI: 1.005–1.008), being female (OR: 1.19, 95% CI: 1.13–1.30), and rural residence (OR: 2.04, 95% CI: 1.93–2.15). To include the province of residence in this model, the provinces of the country were first ranked from large to small based on their non-response rates, then they were given a rank from 1 to 31. Thereafter, the rank of each province based on the non-response rate was entered into the model. The OR of the province of residence indicated the rise in the chance of responding with each increase in the rank of the non-response rate of the province.

**Table 1:** Demographic characteristics of the participants by province.

|  | Sample size | No. of participants (%) | Age, years |  |  |  |  | Gender |  | Area |  |
| --- | --- | --- | --- | --- | --- | --- | --- | --- | --- | --- | --- |
|  |  |  | Mean (SD) | 6-17<sup>a</sup> | 18-39<sup>a</sup> | 40-59<sup>a</sup> | 60≥<sup>a</sup> | Male<sup>a</sup> | Female<sup>a</sup> | Urban<sup>a</sup> | Rural<sup>a</sup> |
| Country Total | 28314 | 11256 | 35.9 | 2302 | 4496 | 3100 | 1358 | 5406 | 5850 | 6851 | 4405 |
|  |  | (39.7) | (18.6) | (20.45) | (39.94) | (27.54) | (12.06) | (48.03) | (51.97) | (60.86) | (39.13) |
| Alborz | 858 | 231 | 35.6 | 44 | 99 | 66 | 22 | 98 | 133 | 215 | 16 |
|  |  | (26.9) | (16.9) | (19) | (42.9) | (28.6) | (9.5) | (42.4) | (57.6) | (93.1) | (6.9) |
| Ardabil | 858 | 223 | 37.4 | 45 | 81 | 69 | 28 | 116 | 107 | 148 | 75 |
|  |  | (26.0) | (18.6) | (20.2) | (36.3) | (30.9) | (12.6) | (52) | (48) | (66.4) | (33.6) |
| Bushehr | 858 | 346 | 36.1 | 65 | 140 | 108 | 33 | 169 | 177 | 246 | 100 |
|  |  | (40.3) | (17.4) | (18.8) | (40.5) | (31.2) | (9.5) | (48.8) | (51.2) | (71.1) | (28.9) |
| Chaharmahal & Bakhtiari | 858 | 310 | 36.4 | 64 | 113 | 93 | 40 | 156 | 154 | 205 | 105 |
|  |  | (36.1) | (18.7) | (20.6) | (36.5) | (30) | (12.9) | (50.3) | (49.7) | (66.1) | (33.9) |
| East Azerbaijan | 858 | 210 | 36.9 | 39 | 80 | 62 | 29 | 99 | 111 | 119 | 91 |
|  |  | (24.5) | (18.9) | (18.6) | (38.1) | (29.5) | (13.8) | (47.1) | (52.9) | (56.7) | (43.3) |
| Fars | 858 | 271 | 36.9 | 53 | 103 | 74 | 41 | 129 | 142 | 159 | 112 |
|  |  | (31.6) | (19.2) | (19.6) | (38) | (27.3) | (15.1) | (47.6) | (52.4) | (58.7) | (41.3) |
| Guilan | 858 | 218 | 42.1 | 30 | 68 | 79 | 41 | 100 | 118 | 109 | 109 |
|  |  | (25.4) | (19.2) | (13.8) | (31.2) | (36.2) | (18.8) | (45.9) | (54.1) | (50) | (50) |
| Golestan | 858 | 362 | 33.8 | 95 | 140 | 89 | 38 | 167 | 195 | 127 | 235 |
|  |  | (42.2) | (18.5) | (26.2) | (38.7) | (24.6) | (10.5) | (46.1) | (53.9) | (35.1) | (64.9) |
| Hamadan | 858 | 202 | 38.1 | 42 | 72 | 53 | 35 | 98 | 104 | 120 | 82 |
|  |  | (23.5) | (20.1) | (20.8) | (35.6) | (26.2) | (17.3) | (48.5) | (51.5) | (59.4) | (40.6) |
| Hormozgan | 858 | 336 | 34.7 | 72 | 141 | 84 | 39 | 151 | 185 | 108 | 228 |
|  |  | (39.2) | (18.9) | (21.4) | (42) | (25) | (11.6) | (44.9) | (55.1) | (32.1) | (67.9) |
| Ilam | 858 | 360 | 35.1 | 73 | 159 | 82 | 46 | 186 | 174 | 217 | 143 |
|  |  | (42.0) | (18.6) | (20.3) | (44.2) | (22.8) | (12.8) | (51.7) | (48.3) | (60.3) | (39.7) |
| Isfahan | 858 | 368 | 39.6 | 58 | 132 | 112 | 66 | 184 | 184 | 302 | 66 |
|  |  | (42.9) | (19.4) | (15.8) | (35.9) | (30.4) | (17.9) | (50) | (50) | (82.1) | (17.9) |
| Kerman | 858 | 474 | 34.0 | 126 | 186 | 108 | 54 | 251 | 223 | 250 | 224 |
|  |  | (55.2) | (19.3) | (26.6) | (39.2) | (22.8) | (11.4) | (53) | (47) | (52.7) | (47.3) |
| Kermanshah | 858 | 349 | 37.0 | 60 | 139 | 81 | 50 | 151 | 179 | 187 | 143 |
|  |  | (40.7) | (19.8) | (18.2) | (42.1) | (24.5) | (15.2) | (45.8) | (54.2) | (56.7) | (43.3) |
| Khuzestan | 858 | 416 | 34.1 | 91 | 183 | 103 | 39 | 196 | 220 | 284 | 132 |
|  |  | (48.5) | (17.8) | (21.9) | (44) | (24.8) | (9.4) | (47.1) | (52.9) | (68.3) | (31.7) |
| Kohgiluyeh & Boyer-Ahmad | 858 | 321 | 34.4 | 70 | 132 | 90 | 29 | 155 | 166 | 168 | 153 |
|  |  | (37.4) | (17.5) | (21.8) | (41.1) | (28) | (9) | (48.3) | (51.7) | (52.3) | (47.7) |
| Kurdistan | 858 | 429 | 37.7 | 82 | 158 | 124 | 65 | 199 | 230 | 263 | 166 |
|  |  | (50.0) | (19.4) | (19.1) | (36.8) | (28.9) | (15.2) | (46.4) | (53.6) | (61.3) | (38.7) |
| Lorestan | 858 | 265 | 33.9 | 72 | 101 | 63 | 29 | 141 | 124 | 140 | 125 |
|  |  | (30.9) | (19.1) | (27.2) | (38.1) | (23.8) | (10.9) | (53.2) | (46.8) | (52.8) | (47.2) |
| Markazi | 858 | 340 | 40.9 | 52 | 109 | 118 | 61 | 156 | 184 | 234 | 106 |
|  |  | (39.6) | (19.4) | (15.3) | (32.1) | (34.7) | (17.9) | (45.9) | (54.1) | (68.8) | (31.2) |
| Mazandaran | 858 | 569 | 38.8 | 89 | 229 | 169 | 82 | 275 | 294 | 290 | 279 |
|  |  | (66.3) | (18.7) | (15.6) | (40.2) | (29.7) | (14.4) | (48.3) | (51.7) | (51) | (49) |
| North Khorasan | 858 | 464 | 34.5 | 103 | 187 | 124 | 50 | 218 | 246 | 217 | 247 |
|  |  | (54.1) | (19.0) | (22.2) | (40.3) | (26.7) | (10.8) | (47) | (53) | (46.8) | (53.2) |
| Qazvin | 858 | 334 | 36.5 | 55 | 142 | 104 | 33 | 159 | 175 | 221 | 113 |
|  |  | (38.9) | (17.3) | (16.5) | (42.5) | (31.1) | (9.9) | (47.6) | (52.4) | (66.2) | (33.8) |
| Qom | 858 | 198 | 34.3 | 43 | 78 | 63 | 14 | 94 | 104 | 177 | 21 |
|  |  | (23.1) | (16.6) | (21.7) | (39.4) | (31.8) | (7.1) | (47.5) | (52.5) | (89.4) | (10.6) |
| Razavi Khorasan | 858 | 732 | 37.5 | 95 | 332 | 220 | 85 | 375 | 357 | 555 | 177 |
|  |  | (85.3) | (16.7) | (13) | (45.5) | (30.1) | (11.6) | (51.2) | (48.8) | (75.8) | (24.2) |
| Semnan | 858 | 359 | 39.1 | 59 | 131 | 112 | 57 | 173 | 186 | 281 | 78 |
|  |  | (41.8) | (19.1) | (16.4) | (36.5) | (31.2) | (15.9) | (48.2) | (51.8) | (78.3) | (21.7) |
| Sistan & Baluchestan | 858 | 650 | 29.1 | 207 | 287 | 117 | 39 | 302 | 348 | 264 | 386 |
|  |  | (75.8) | (17.4) | (31.8) | (44.2) | (18) | (6) | (46.5) | (53.5) | (40.6) | (59.4) |
| South Khorasan | 858 | 390 | 33.7 | 106 | 144 | 101 | 39 | 172 | 218 | 209 | 181 |
|  |  | (45.4) | (19.0) | (27.2) | (36.9) | (25.9) | (10) | (44.1) | (55.9) | (53.6) | (46.4) |
| Tehran | 2574 | 248 | 36.4 | 41 | 108 | 81 | 18 | 116 | 132 | 220 | 28 |
|  |  | (9.6) | (16.2) | (16.5) | (43.5) | (32.7) | (7.3) | (46.8) | (53.2) | (88.7) | (11.3) |
| West Azerbaijan | 858 | 520 | 34.6 | 115 | 204 | 141 | 60 | 258 | 262 | 316 | 204 |
|  |  | (60.6) | (18.7) | (22.1) | (39.2) | (27.1) | (11.5) | (49.6) | (50.4) | (60.8) | (39.2) |
| Yazd | 858 | 296 | 34.5 | 68 | 115 | 83 | 30 | 143 | 153 | 234 | 62 |
|  |  | (34.5) | (18.0) | (23) | (38.9) | (28) | (10.1) | (48.3) | (51.7) | (79.1) | (20.9) |
| Zanjan | 858 | 484 | 36.4 | 88 | 203 | 127 | 66 | 219 | 265 | 266 | 218 |
|  |  | (56.4) | (18.9) | (18.2) | (41.9) | (26.2) | (13.6) | (45.2) | (54.8) | (55) | (45) |

a: Number (%)

**Figure 1:**
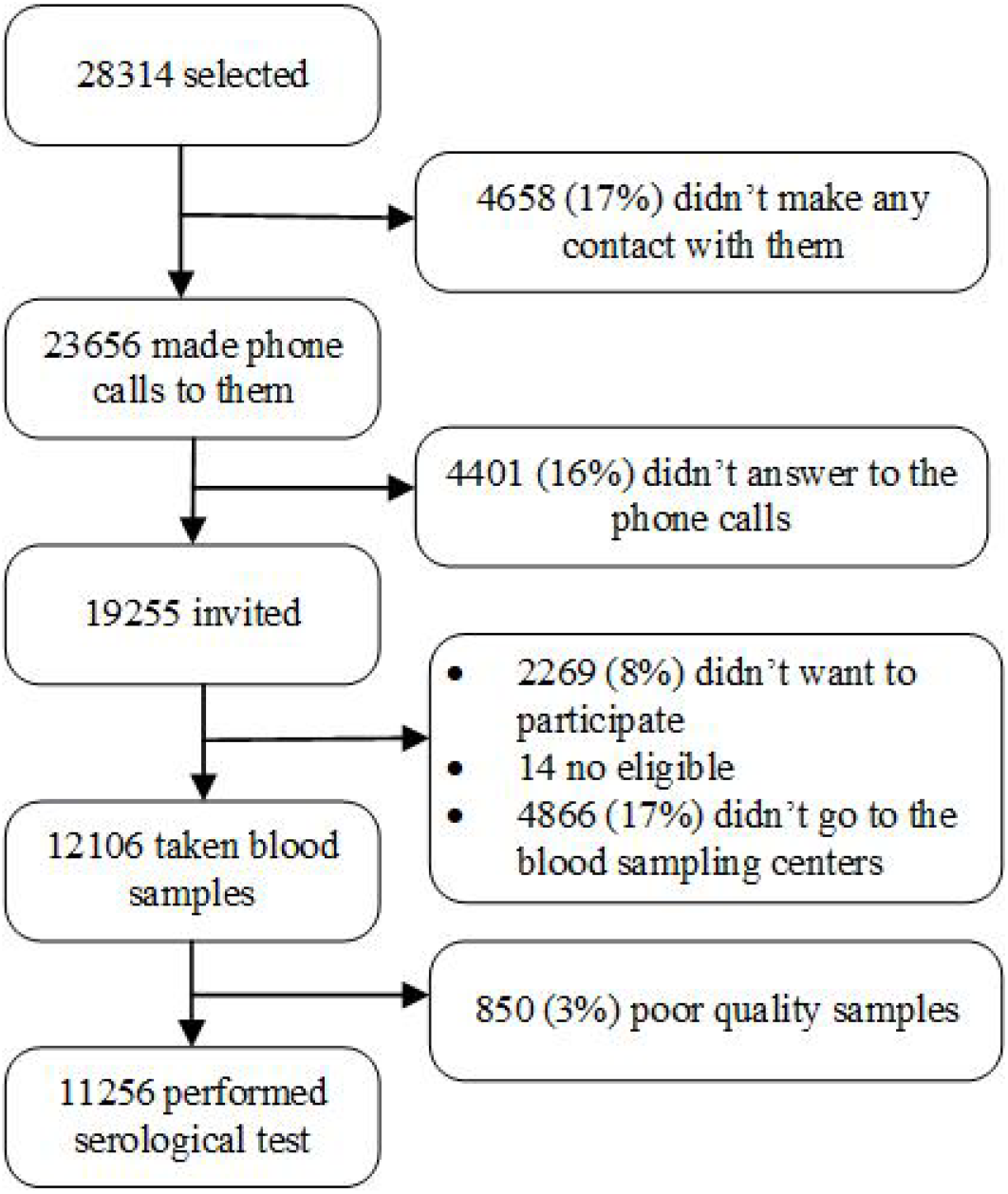
Profile of participation in the study.

The blood sampling of the study participants started on August 3, 2020, in Tehran province and continued until October 31, 2020, in Kohgiluyeh and Boyer-Ahmad province. Half the participants were sampled before September 10 and the other half after that date. To increase the response rate, we extended the data collection period by one month. The mean (standard deviation) of the age of the study participants in the country was 35.89 (18.61) years (age range, 6–109 years). In total, 5406 (48.03%) of the participants were male, and 6851 (60.86%) lived in urban areas. Table 1 shows the distribution of the age, sex, and area of residence of the study participants across the country and by province.

We estimated the sensitivity and specificity of the IgG test of the study laboratory kit as 0.74 (95% CI: 0.67, 0.80) and 0.98 (95% CI: 0.96, 0.99), respectively. Further details on the characteristics of the participants in the sensitivity and specificity estimation studies and their results are provided in Appendix B.

Of the 11,256 participants in the study nationally, the IgG serological test was positive in 1303 (11.6%, 95% CI: 11.0, 12.0). The crude prevalence of COVID-19 based on the IgG serological testing is presented in total in Figure 2 and separately by age, sex, and urban/rural subgroup in Table S3 of Appendix C for the country and the provinces. The prevalence estimates with a 95% CI wider than 12% have not been reported in Table S3 due to low precision.

**Figure 2:**
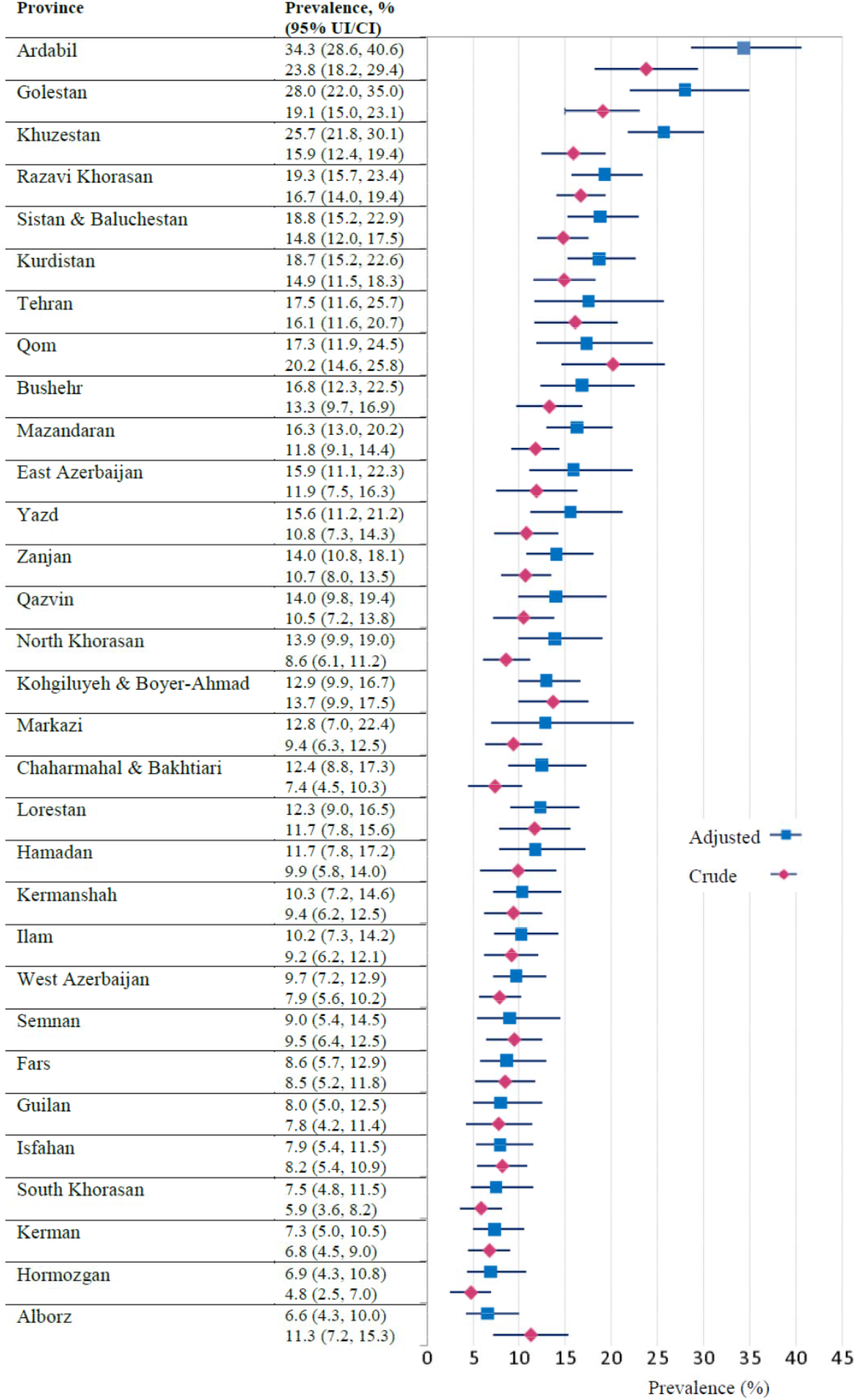
The crude (unadjusted) prevalence (95% CI) and test measurement error adjusted and weighted prevalence (95% UI) of COVID-19 in Iran (by province)

The prevalence of COVID-19 in Iran after correcting for the laboratory kit measurement error and weighting data by post-stratification, inverse probability of response, and sampling design weights was estimated as 14.2% (95% UI: 13.3, 15.2) as at August 20, 2020. It was therefore estimated that from the beginning of the pandemic to this date, more than 11,958,346 (95% CI: 11,211,011–12,746,776) of the population over six years of age had been infected with COVID-19 in Iran. Nationwide, the infection prevalence was higher among men than women, urban than rural populations, and individuals ≥60 years of age than other age groups (Table S3 of Appendix C). The corrected prevalence estimates by age, gender, and urban/rural area of residence for each province are also presented in Table S3; the prevalence estimates with a 95% UI wider than 12% are not reported in this table. Ardabil, Golestan, and Khuzestan provinces had the highest prevalence, and Alborz, Hormozgan, and Kerman provinces had the lowest (see Figures 2 and 3).

**Figure 3:**
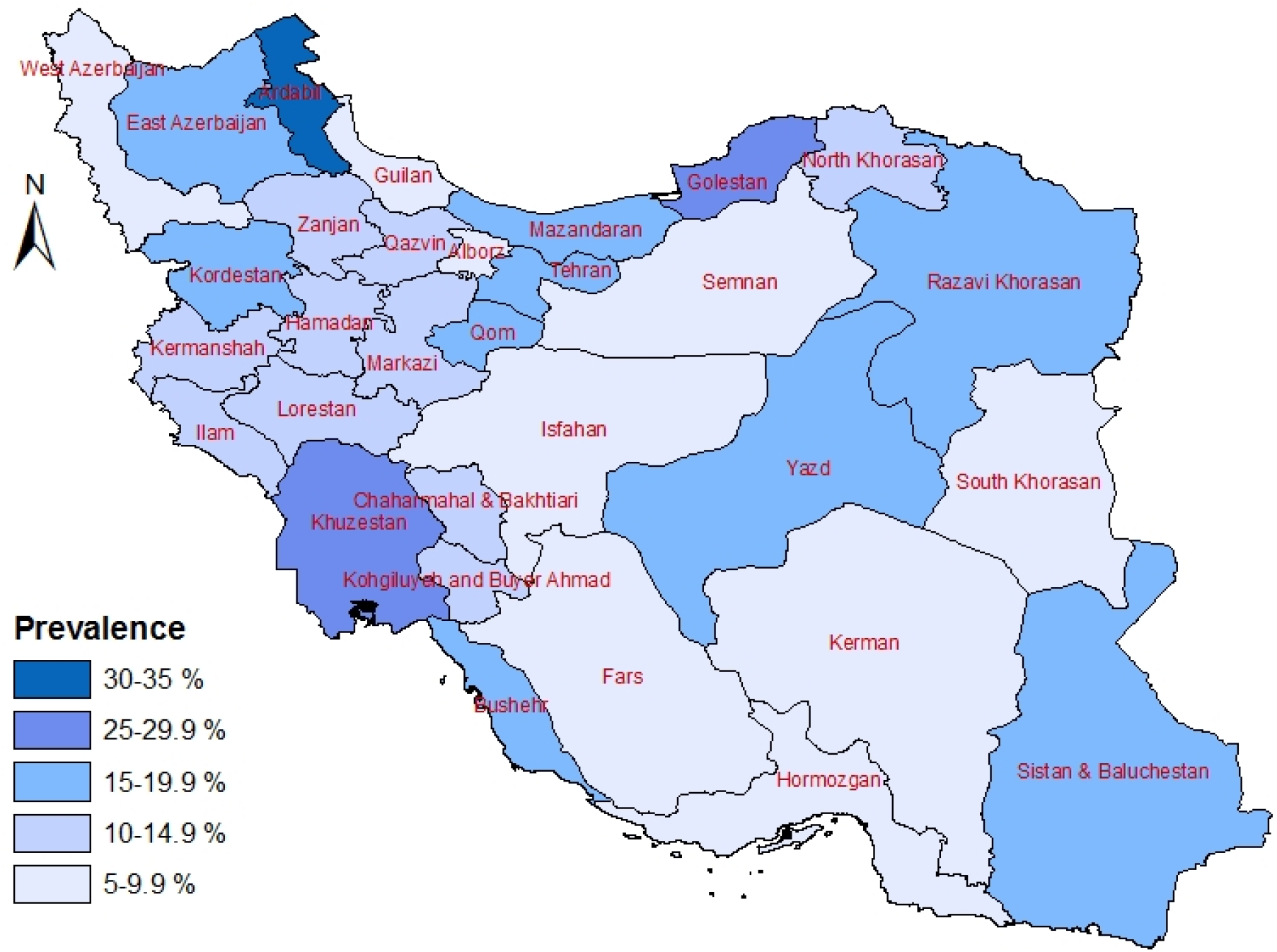
Spatial distribution of the COVID-19 in Iran (test measurement error adjusted and weighted prevalence of the infection until August 20, 2020)

Figure 2 presents the crude prevalence estimates and the prevalence estimates corrected for the laboratory kit measurement error and weighting by all weights among the provinces.

## Discussion

We estimated the prevalence of COVID-19 in the population aged six years and older in Iran from the beginning of the pandemic until August 20, 2020, as 14.2%. Our study showed that the prevalence of the infection was higher among men than women, in the urban than the rural population, and in the age groups ≥60 years and 40–59 years than the other age groups. Ardabil, Golestan, Khuzestan, Razavi Khorasan, and Sistan and Baluchestan provinces had the highest prevalence, and Alborz, Hormozgan, Kerman, South Khorasan, and Isfahan provinces had the lowest.

In a study conducted in Iran on the general population in 18 of the most densely populated metropolises across 17 provinces in the country, the prevalence of COVID-19 from the start of the pandemic until the end of April 2020 was estimated as 17.1% (11, 13-15). Unlike our study, this study did not capture the data of patients four months after April, so the estimated prevalence was higher than our national prevalence estimate. The estimated prevalence of the disease would likely have been higher than that in our study because the sampling population of the study only included metropolises with high population densities—not small and sparsely populated cities and villages—and the study was conducted only in provinces with the highest reported number of COVID-19 cases based on MOHME reports (13). Although the laboratory kit used in the study was the same as ours, the sensitivity of the IgG test was estimated in different situations (within 2–4 weeks of symptom onset vs. 3–16 weeks in our study) and was found to be lower than that in our study (61% vs. 74%, respectively). In terms of COVID-19 infection, the IgG titer increased after week 3 of symptom onset, but the samples were used within 2–4 weeks in their sensitivity estimation study. These factors, as well as some of the study’s methodological and statistical issues (15), may have contributed to the differences between the results of that study and ours.

In another seroprevalence study in Guilan province, the prevalence of COVID-19 from the beginning of the pandemic until April 2020 was estimated at 22.2% (12), which is significantly higher than our estimate for Guilan province (8.0%). There could be several reasons for this discrepancy, including household sampling compared to the simple random sampling undertaken in our study (due to the high risk of infection of all household members if one of the members is infected), the inclusion of only a number of districts with high and low incidences (based on their hospitalization rate) compared to the inclusion of all districts in the present study, and possibly greater participation of infected subjects in that study compared to ours due to subjects’ fear of becoming infected in the comprehensive health centers (because of this, 17% of the subjects did not participate in that study). More importantly, as Guilan province was among the first provinces hit by the COVID-19 pandemic, it is quite possible that the proportion of infected people with negative serologic test results would have been higher than that of many other provinces with delayed peaks. This may have led to a higher underestimation of the infection in Guilan province in our study. In terms of statistical methods, the principle of order in the multiple bias correction in that study was disregarded so that the measurement error correction could be done before the nonresponse bias correction with inverse probability weighting, 27) (28. This could also have affected the results. Additionally, not using weighting within the strata of the age, gender, and rural/urban variables in the Guilan study could have resulted in differences. These factors may also be the reason for the different age, sex, and urban/rural distribution of the disease in that study compared to ours.

In our study, the prevalence of COVID-19 infection at school age (6–17 years) was estimated at 11.5%. In the studies of Poustchi et al. (11) and Shakiba et al. (12), the prevalence of infection in this age group was also high (14.3% and 19.1%, respectively). On the one hand, a high proportion of the patients in this age group were asymptomatic or had mild symptoms (29), and on the other hand, this population may play an important role in the spread of the infection (30) because hygiene and the principles of personal protection, especially in younger age groups, are weak (31). Therefore, to control the pandemic, it is necessary to take special measures to limit their presence in society through remote education or face-to-face education only in the case of older age groups, with strict observance of social distancing, proper classroom ventilation, the continuous use of masks, and frequent hand washing and disinfection.

The increase in the number of definitive cases identified and the daily deaths due to COVID-19 have created the misconception that a large proportion of the population may already have been infected. However, to date, the results of population-based seroprevalence studies in most parts of the world have not confirmed this hypothesis (33, 32, 9). In Iran, the increase in the number of daily deaths along with the results of two previous seroprevalence studies (11, 12) have created this perspective among policymakers and the public; hence, the results of our study have the potential to correct this misconception.

Our study had some limitations. The first was the use of populations registered in the PHC electronic health record systems as a sampling framework. Despite the coverage of over 90% of the population of many provinces of the country by these systems, in some provinces, such as Tehran province, the coverage of this system was about 80%. When calculating the prevalence estimates by weighting, the distribution of age, sex, urban/rural, and province of residence of the survey sample was matched with the distribution of the provinces and the country population (based on the projection of the population in 2020 by the Statistics Center of Iran). Accordingly, when estimating the COVID-19 prevalence, the problem of the incomplete coverage of the sampling framework was partially solved, with the exception of the influential variables not included in our weightings, such as the distribution of socioeconomic status. Second, there was a high rate of non-responses in our study. This reduced the precision of the estimates and increased the risk of selection bias. To counter the possibility of selection bias, the elements of such bias relating to gender, age, urban/rural location, and province of residence due to non-responses were modified during the statistical analysis by inverse probability of response weighting within the joint categories of these variables (496 categories). Given the association between these variables and several other factors that may have caused non-responses, it is expected that the weighting controlled most of the non-response bias of the present survey. Notwithstanding, we were not able to control for a small amount of selection bias because we did not know the status of the other variables affecting the non-responses, including the distribution of severe cases, socioeconomic status, effective isolation, and contact tracing in the different provinces. Third, in the laboratory kit sensitivity study, the included patients were individuals who presented at 3–16 weeks following disease symptom onset. This combination of patients did not include those individuals who presented after 16 weeks from symptom onset. Such subjects may have provided false-negative results due to a drop in the antibody titers following the extended period since the onset of the symptoms. We may therefore have estimated the sensitivity of the study kit as being slightly higher than the true value. It should be noted, however, that when estimating the final corrected prevalence in our study using this estimated sensitivity, all the false-negative results of the patients who presented less than four months after the onset of symptoms had been corrected. We therefore maintain that we identified all patients with the infection from May 10 to August 20, 2020, in addition to those before May 10 who had the IgG antibody at the time of the survey (the median of the sampling duration of our survey was September 10, 2020). Accordingly, the true prevalence estimates from the start of the pandemic to August 20, 2020, are expected to be slightly higher than the estimates in this survey, especially for the provinces with earlier peaks in the pandemic.

## Conclusion

To the best of our knowledge, this study is the only nationwide population-based survey in Iran and among only a few in the world. One of the strengths of this survey is its simultaneous correction of both the measurement error of the laboratory kit and the non-response bias. Despite the stated limitations when interpreting the results, the results of this survey provide a clear picture of the status of COVID-19 in the general population of Iran at a national and provincial level for use in policy-making and planning. According to the study results, a large proportion of the population had not yet been infected. The observance of the principles of hygiene and social restrictions should therefore continue until the majority of the population has been vaccinated. The experience of this survey led us to plan the next ICS program surveys with the aim of attaining a higher response rate and estimating the sensitivity of the study kit more accurately by performing serological testing on a number of positive PCR patients from early in the pandemic to mid-May 2020.

## Data Availability

All data referred to in the manuscript are not available.

## Acknowledgements

This study, the first survey of the ICS program, was ordered and funded by the Deputy of Public Health of the Iranian MOHME and the National Institute for Health Research. It was implemented through the active participation of the Network Management Center, the National Comprehensive Health Laboratory, and the Centers for Communicable and Non-Communicable Diseases Control of the MOHME, as well as the departments of health laboratories, communicable diseases, and network management at medical universities, and district primary health networks across the country. We are grateful for the cooperation of the staff of the comprehensive health centers, rural health houses, urban health posts, and health laboratories throughout the country.

## References

1. Zhu N, Zhang D, Wang W, Li X, Yang B, Song J, et al. A novel coronavirus from patients with pneumonia in China, 2019. New England Journal of Medicine. 2020.

2. Hu B, Guo H, Zhou P, Shi Z-L. Characteristics of SARS-CoV-2 and COVID-19. Nature Reviews Microbiology. 2020:1–14.

3. WHO. Population-based age-stratified seroepidemiological investigation protocol for COVID-19 virus infection, 17 March 2020. World Health Organization; 2020.

4. Christoph H. Coronavirus Update: Islamic Repulic of Iran, Coronavirus Disease 2019 (COVID-19), 01 December 2020. WHO Iran Country Office, Tehran; 2020.

5. Arevalo-Rodriguez I, Buitrago-Garcia D, Simancas-Racines D, Zambrano-Achig P, Del Campo R, Ciapponi A, et al. False-negative results of initial RT-PCR assays for COVID-19: a systematic review. PloS one. 2020;15(12):e0242958.

6. Byambasuren O, Cardona M, Bell K, Clark J, McLaws M-L, Glasziou P. Estimating the extent of asymptomatic COVID-19 and its potential for community transmission: systematic review and meta-analysis. Official Journal of the Association of Medical Microbiology and Infectious Disease Canada. 2020;5(4):223–34.

7. He J, Guo Y, Mao R, Zhang J. Proportion of asymptomatic coronavirus disease 2019: A systematic review and meta-analysis. Journal of medical virology. 2021;93(2):820–30.

8. Bryant JE, Azman AS, Ferrari MJ, Arnold BF, Boni MF, Boum Y, et al. Serology for SARS-CoV-2: apprehensions, opportunities, and the path forward. Science Immunology. 2020;5(47).

9. Pollán M, Pérez-Gómez B, Pastor-Barriuso R, Oteo J, Hernán MA, Pérez-Olmeda M, et al. Prevalence of SARS-CoV-2 in Spain (ENE-COVID): a nationwide, population-based seroepidemiological study. The Lancet. 2020;396(10250):535–44.

10. Theel ES, Slev P, Wheeler S, Couturier MR, Wong SJ, Kadkhoda K. The role of antibody testing for SARS-CoV-2: is there one? Journal of clinical microbiology. 2020;58(8).

11. Poustchi H, Darvishian M, Mohammadi Z, Shayanrad A, Delavari A, Bahadorimonfared A, et al. SARS-CoV-2 antibody seroprevalence in the general population and high-risk occupational groups across 18 cities in Iran: a population-based cross-sectional study. The Lancet Infectious Diseases. 2020.

12. Shakiba M, Nazemipour M, Salari A, Mehrabian F, Nazari SS, Rezvani SM, et al. Seroprevalence of SARS-CoV-2 in Guilan Province, Iran, April 2020. Emerging Infectious Diseases. 2020;27(2):636–8.

13. Darvishian M, Sharafkhah M, Poustchi H, Malekzadeh R. Estimates of anti-SARS-CoV-2 antibody seroprevalence in Iran – Authors’ reply. The Lancet Infectious Diseases. 2021.

14. Ghafari M, Kadivar A, Katzourakis A. Estimates of anti-SARS-CoV-2 antibody seroprevalence in Iran. The Lancet Infectious Diseases. 2021.

15. Nazemipour M, Shakiba M, Mansournia MA. Estimates of anti-SARS-CoV-2 antibody seroprevalence in Iran. The Lancet Infectious Diseases. 2021.

16. Khalagi K, Gharibzadeh S, Mirab Samiee S, Hashemi SM, Khalili D, Aghamohamadi S, et al. Nationwide population-based surveys of Iranian COVID-19 Serological Surveillance (ICS) program: The surveys protocol. Med J Islam Repub Iran. 2021;35(61).

17. Shakiba M, Nazari SSH, Mehrabian F, Rezvani SM, Ghasempour Z, Heidarzadeh A. Seroprevalence of COVID-19 virus infection in Guilan province, Iran. medRxiv. 2020.

18. Deeks JJ, Dinnes J, Takwoingi Y, Davenport C, Spijker R, Taylor-Phillips S, et al. Antibody tests for identification of current and past infection with SARS-CoV-2. Cochrane Database of Systematic Reviews. 2020(6).

19. To KK-W, Tsang OT-Y, Leung W-S, Tam AR, Wu T-C, Lung DC, et al. Temporal profiles of viral load in posterior oropharyngeal saliva samples and serum antibody responses during infection by SARS-CoV-2: an observational cohort study. The Lancet Infectious Diseases. 2020.

20. Azizi F, Takyar M, Zadeh-Vakili A. Contributions and Implications of the Tehran Lipid and Glucose Study. Int J Endocrinol Metab. 2018;16(4 Suppl):e84792.

21. Zou G. A Modified Poisson Regression Approach to Prospective Studies with Binary Data. American Journal of Epidemiology. 2004;159(7):702–6.

22. Diggle PJ. Estimating prevalence using an imperfect test. Epidemiology Research International. 2011;2011.

23. Mansournia MA, Altman DG. Inverse probability weighting. Bmj. 2016;352.

24. Mansournia MA, Nazemipour M, Naimi AI, Collins GS, Campbell MJ. Reflections on modern methods: demystifying robust standard errors for epidemiologists. International Journal of Epidemiology. 2020.

25. StataCorp L. Stata Statistical Software: Release 15 College Station, TX. StataCorp; 2017.

26. Team RC. R: A language and environment for statistical computing. R Foundation for Statistical Computing, Vienna, Austria2020.

27. Lash TL, Fox MP, Fink AK. Applying quantitative bias analysis to epidemiologic data: Springer Science & Business Media; 2011.

28. Rothman KJ, Greenland S, Lash TL. Modern epidemiology: Lippincott Williams & Wilkins; 2008.

29. Chang T-H, Wu J-L, Chang L-Y. Clinical characteristics and diagnostic challenges of pediatric COVID-19: A systematic review and meta-analysis. Journal of the Formosan Medical Association. 2020.

30. Hyde Z. COVID-19, children, and schools: overlooked and at risk. Med J Aust. 2020;213(10):444–6.

31. Stage HB, Shingleton J, Ghosh S, Scarabel F, Pellis L, Finnie T. Shut and re-open: the role of schools in the spread of COVID-19 in Europe. arXiv preprint arXiv:200614158. 2020.

32. Bajema KL, Wiegand RE, Cuffe K, Patel SV, Iachan R, Lim T, et al. Estimated SARS-CoV-2 Seroprevalence in the US as of September 2020. JAMA internal medicine. 2020.

33. Hallal PC, Hartwig FP, Horta BL, Silveira MF, Struchiner CJ, Vidaletti LP, et al. SARS-CoV-2 antibody prevalence in Brazil: results from two successive nationwide serological household surveys. The Lancet Global Health. 2020;8(11):e1390–e8.

