## Supplementary Appendix for "Prevalence of COVID-19 in Iran: Results of the first survey of the Iranian COVID-19 Serological Surveillance program"

#### **Appendix A:** The survey implementation process

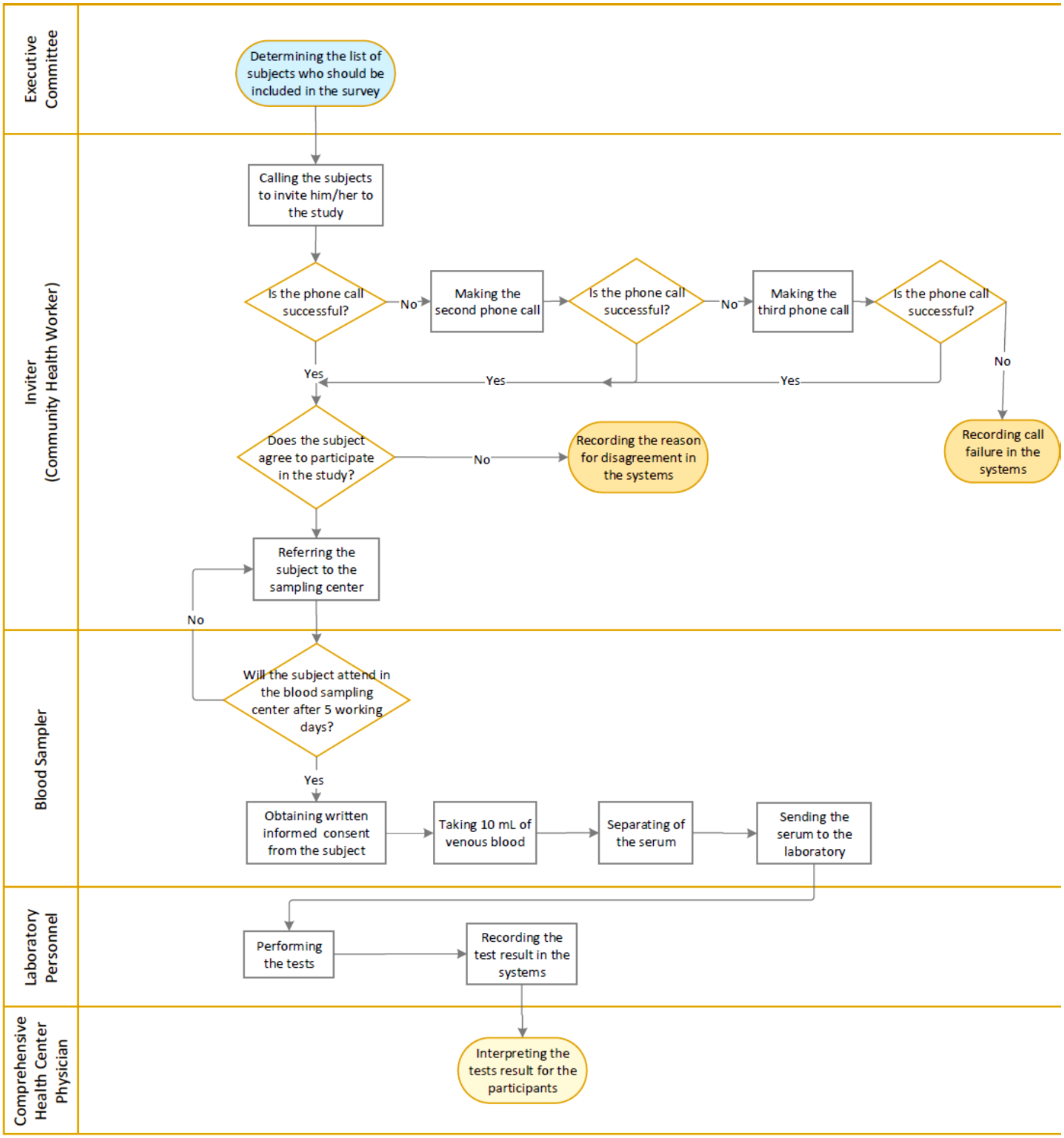


Figure S1: The survey implementation process

**Appendix B:** Estimating the sensitivity and specificity of Pishtaz Teb laboratory kit

In order to estimate the sensitivity of Pishtaz Teb laboratory kit in comparison with the gold standard PCR test in the diagnosis of COVID- 19, blood samples were taken from 254 cases with a definite diagnosis of COVID-19 based on the PCR test, and IgG antibodies were tested in their serum using the study laboratory kit. Table S1 presents the age and sex distributions, the time elapsed since the onset of symptoms, and hospitalization status of the subjects included in the sensitivity calculation study.

Table S1: Characteristics of the participants in the study of estimating the sensitivity of the Pishtaz-Teb kit

| | Female *No. (%)* | Male *No. (%)* | Total *No. (%)* |
|---|---|---|---|
| ***Age (years)*** | | | |
| 6-20 | 6 (4.41) | 3 (2.54) | 9 (3.54) |
| 21-30 | 13 (9.56) | 11 (9.32) | 24 (9.45) |
| 31-40 | 38 (27.94) | 28 (23.73) | 66 (25.98) |
| 41-50 | 25 (18.38) | 26 (22.03) | 51 (20.08) |
| 51-60 | 27 (19.85) | 24 (20.34) | 51 (20.08) |
| ≥ 60 | 27 (19.85) | 26 (22.03) | 53 (20.87) |
| ***Interval from onset of the symptoms (week)*** | | | |
| 3-4 | 41 (30.15) | 35 (29.66) | 76 (29.92) |
| 5-8 | 45 (33.09) | 36 (30.51) | 81 (31.89) |
| 9-12 | 43 (31.62) | 33 (27.97) | 76 (29.92) |
| 13-16 | 7 (5.15) | 14 (11.86) | 21 (8.27) |
| ***Hospitalization status*** | | | |
| Outpatient | 85 (62.50) | 56 (47.46) | 141 (55.51) |
| Inpatient | 51 (37.50) | 62 (52.54) | 113 (44.49) |

Evaluation of the factors affecting the positive result of IgG serology test in the PCR positive subjects (sensitivity) by multiple modified Poisson regression showed no evidence for association between sensitivity with age (P=0.7), sex (P=0.47), severity of symptoms (inpatient/ outpatient care)) (P=0.29), and duration after symptom onset (week) (P=0.11). The modified Poisson regression gave us the relative risk (RR) of sensitivity for each of the mentioned independent variables (1). On the other hand, the one-way chi-square test showed a little evidence for association between severity of symptoms and sensitivity of the IgG test (P=0.08). Accordingly, the sensitivity of the IgG test of the study laboratory kit after weighting based on the distribution of severity of disease symptoms in the community (assuming that 10% of the patients had severe symptoms and 90% had no severe symptoms) was estimated as 0.74; 95% CI: (0.67, 0.80).

To estimate the specificity of Pishtaz Teb laboratory kit, frozen serum of 410 participants in the Tehran Lipid and Glucose Study (TLGS) (2) was used, and the presence of IgG antibody against COVID-19 in their serum was tested using the study laboratory kit. Table S2 shows the age and sex distributions and the season when serum samples were taken from these individuals. Investigating the factors affecting the positive serological test result of IgG (false positive or test specificity) using multiple modified Poisson regression in healthy individuals showed that age is the only factor that affects the positive serological test result in healthy individuals. So that, increasing in age is associated with an increase in false-positive probability (RR = 1.03 (95%CI: 1.00, 1.05)), and a decrease in test specificity. There was no evidence for association of gender (P=0.12) and season of blood sampling (P>0.2) with the test specificity. Therefore, the specificity of IgG serology test for the study laboratory kit, after weighting based on the age distribution of the Iranian population in the 2016 census, was estimated as 0.98 (95%CI: 0.96, 0.99).

Table S2: Characteristics of the subjects whose frozen serum was used to estimate the specificity of the Pishtaz-Teb kit

| | Female<br>*No. (%)* | Male<br>*No. (%)* | Total<br>*No. (%)* |
|---|---|---|---|
| ***Age (years)*** | | | |
| 6-20 | 37 (18.14) | 33 (16.02) | 70 (17.07) |
| 21-30 | 36 (17.65) | 46 (22.33) | 82 (20) |
| 31-40 | 56 (27.45) | 53 (25.73) | 109 (26.59) |
| 41-50 | 28 (13.73) | 24 (11.65) | 52 (12.68) |
| 51-60 | 24 (11.76) | 28 (13.59) | 52 (12.68) |
| ≥ 60 | 23 (11.27) | 22 (10.68) | 45 (10.98) |
| ***Sampling season*** | | | |
| Spring | 51 (25) | 52 (25.24) | 103 (25.12) |
| Summer | 52 (25.49) | 51 (24.76) | 103 (25.12) |
| Fall | 51 (25) | 52 (25.24) | 103 (25.12) |
| Winter | 50 (24.51) | 51 (24.76) | 101 (24.63) |

**Appendix C:** Prevalence of COVID-19 in Iran and among the provinces by age, gender and urban/rural area of residence

Table S3: Crude prevalence vs. test measurement error adjusted and weighted prevalence of COVID-19 in Iran and among the provinces (by age, gender and urban/rural area of residence)

| | No. of participants | No. of IgG positives | Prevalence (95% CI/UI) [a] | Gender | | Area | | Age, years | | | |
|---|---|---|---|---|---|---|---|---|---|---|---|
| | | | | Male | Female | Urban | Rural | 6-17 | 18-39 | 40-59 | 60≥ |
| Total Country | 11256 | 1303 | Crude | 11.3 (10.5, 12.2) | 11.8 (11.0, 12.6) | 12.4 (11.6, 13.2) | 10.3 (9.4, 11.2) | 10.3 (9.1, 11.5) | 10.3 (9.5, 11.2) | 13.0 (11.8, 14.2) | 14.7 (12.8, 16.5) |
| | | | Adjusted | 14.6 (13.4, 15.9) | 13.8 (12.5, 15.2) | 16.6 (15.3, 18.0) | 11.7 (10.6, 13.0) | 11.5 (9.9, 13.4) | 11.6 (10.5, 12.8) | 14.3 (12.8, 16.0) | 19.4 (17.0, 22.1) |
| Khuzestan | 416 | 67 | Crude | 20.4 (14.8, 26.1) | 11.8 (7.6, 16.1) | 14.8 (10.7, 18.9) | LPE | LPE | 8.2 (4.2, 12.2) | LPE | 15.9 (12.4, 19.4) |
| | | | Adjusted | 35.14 (29.14, 41.64) | 15.8 (11.2, 21.9) | 22.0 (17.0, 27.8) | LPE | LPE | 11.3 (7.2, 17.1) | LPE | LPE |
| Markazi | 340 | 33 | Crude | 7.7 (3.5, 11.9) | 10.9 (6.4, 15.4) | 7.7 (4.3, 11.1) | LPE | LPE | 9.2 (3.8, 14.6) | 11.0 (5.4, 16.7) | LPE |
| | | | Adjusted | 7.4 (3.7, 14.1) | LPE | 7.4 (4.7, 11.4) | LPE | LPE | LPE | LPE | LPE |
| Ardabil | 223 | 53 | Crude | LPE [b] | LPE | LPE | LPE | LPE | LPE | LPE | LPE |
| | | | Adjusted | LPE | LPE | LPE | LPE | LPE | LPE | LPE | LPE |
| West Azerbaijan | 520 | 42 | Crude | 7.0 (3.9, 10.1) | 8.8 (5.4, 12.2) | 8.5 (5.5, 11.6) | 6.9 (3.4, 10.3) | 9.6 (4.2, 14.9) | 4.9 (1.9, 7.9) | 9.2 (4.4, 14.0) | LPE |
| | | | Adjusted | 7.3 (4.4, 11.8) | 11.9 (8.2, 17.0) | 11.6 (8.1, 16.4) | 7.5 (4.4, 12.4) | LPE | 5.3 (2.9, 9.6) | 10.6 (6.4, 17.1) | LPE |
| Hamadan | 202 | 29 | Crude | LPE | 8.7 (3.3, 14.1) | 10.8 (5.3, 16.4) | LPE | LPE | LPE | LPE | LPE |
| | | | Adjusted | LPE | LPE | LPE | LPE | LPE | LPE | LPE | LPE |
| North Khorasan | 464 | 40 | Crude | 8.3 (4.6, 11.9) | 8.9 (5.4, 12.5) | 9.2 (5.4, 13.1) | 8.1 (4.7, 11.5) | 7.8 (2.6, 12.9) | 8.6 (4.5, 12.6) | 8.1 (3.3, 12.9) | LPE |
| | | | Adjusted | LPE | 10.5 (6.8, 15.7) | LPE | 9.0 (6.0, 13.3) | 8.5 (4.4, 15.7) | 10.3 (6.5, 15.8) | 9.8 (5.4, 16.9) | LPE |
| Isfahan | 368 | 31 | Crude | 7.6 (3.8, 11.4) | 8.7 (4.6, 12.8) | 8.3 (5.2, 11.4) | LPE | LPE | 6.1 (2.0, 10.1) | 9.8 (4.3, 15.3) | LPE |
| | | | Adjusted | 5.8 (3.2, 10.4) | 10.0 (6.0, 16.3) | 11.4 (7.6, 16.8) | 4.3 (1.7, 10.7) | LPE | 4.0 (2.1, 7.4) | 8.2 (4.1, 15.7) | LPE |
| Alborz | 231 | 26 | Crude | 9.2 (3.5, 14.9) | 12.8 (7.1, 18.5) | 12.1 (7.7, 16.5) | 0 (0,0) | LPE | LPE | LPE | LPE |

| | | | | | | | | | | | |
|---|---|---|---|---|---|---|---|---|---|---|---|
| | | | Adjusted | 8.7 (4.8, 15.3) | 4.4 (2.6, 7.3) | 20.2 (12.9, 30.1) | 0 (0, 0) | 3.5 (1.6, 7.5) | 4.9 (2.8, 8.4) | 3.2 (1.6, 6.3) | LPE |
| Tehran | 248 | 40 | Crude | LPE | LPE | 16.4 (11.5, 21.3) | LPE | LPE | LPE | LPE | LPE |
| | | | Adjusted | LPE | LPE | LPE | LPE | LPE | LPE | LPE | LPE |
| Sistan & Baluchestan | 650 | 98 | Crude | 13.2 (9.4, 17.1) | 16.1 (12.2, 20.0) | 16.7 (12.2, 21.2) | 13.5 (10.1, 16.9) | 12.6 (8.0, 17.1) | 14.3 (10.2, 18.3) | LPE | LPE |
| | | | Adjusted | LPE | 18.5 (14.4, 23.4) | 19.2 (14.3, 25.2) | 18.2 (13.5, 24.0) | 15.7 (10.7, 22.4) | 17.2 (13.1, 22.1) | LPE | LPE |
| Ilam | 360 | 36 | Crude | 10.2 (5.9, 14.6) | 8.0 (4.0, 12.1) | 10.1 (6.1, 14.2) | 7.7 (3.3, 12.1) | LPE | 11.3 (6.4, 16.2) | 6.1 (0.9, 11.3) | LPE |
| | | | Adjusted | 12.2 (8.2, 17.7) | 8.6 (4.8, 15.0) | 12.0 (7.9, 17.9) | 8.2 (4.6, 14.3) | LPE | LPE | 6.6 (3.1, 13.7) | LPE |
| Mazandaran | 569 | 69 | Crude | 11.6 (7.8, 15.4) | 11.9 (8.2, 15.6) | 13.1 (9.2, 17.0) | 10.4 (6.8, 14.0) | LPE | 9.2 (5.4, 12.9) | 12.4 (7.5, 17.4) | LPE |
| | | | Adjusted | 16.2 (11.7, 22.1) | 16.4 (12.0, 21.9) | 18.7 (13.7, 24.9) | 13.5 (9.7, 18.4) | LPE | 10.7 (7.2, 15.6) | 15.4 (10.6, 21.8) | LPE |
| South Khorasan | 390 | 25 | Crude | 6.4 (2.7, 10.1) | 5.5 (2.5, 8.5) | 6.2 (2.9, 9.5) | 5.5 (2.2, 8.9) | 5.7 (1.3, 10.1) | 4.2 (0.9, 7.4) | 7.9 (2.7, 13.2) | LPE |
| | | | Adjusted | 7.0 (3.7, 12.9) | 8.0 (4.3, 14.6) | 8.3 (4.2, 15.6) | 6.8 (3.8, 12.0) | 5.6 (2.6, 11.6) | 5.2 (2.5, 10.4) | LPE | LPE |
| Kerman | 474 | 32 | Crude | 6.4 (3.4, 9.4) | 7.2 (3.8, 10.6) | 6.4 (3.4, 9.4) | 7.1 (3.8, 10.5) | 7.1 (2.6, 11.6) | 6.5 (2.9, 10.0) | 9.3 (3.8, 14.7) | 1.9 (0, 5.4) |
| | | | Adjusted | 6.6 (4.2, 10.4) | 7.9 (4.5, 13.5) | 5.6 (3.5, 8.8) | 8.9 (5.2, 14.8) | 8.6 (4.7, 15.2) | 7.3 (4.3, 12.2) | 9.8 (5.5, 16.8) | LPE |
| Hormozgan | 336 | 17 | Crude | 6.0 (2.2, 9.7) | 3.8 (1.0, 6.5) | 7.4 (2.5, 12.3) | 3.5 (1.1, 5.9) | 2.8 (0, 6.6) | 5.7 (1.9, 9.5) | 2.4 (0, 5.6) | LPE |
| | | | Adjusted | 8.7 (5.0, 14.7) | 4.8 (2.0, 11.0) | 7.2 (3.8, 13.2) | 6.5 (3.3, 12.4) | LPE | LPE | 2.9 (0.7, 11.0) | LPE |
| Bushehr | 346 | 46 | Crude | 13.0 (7.9, 18.1) | 13.6 (8.5, 18.6) | 13.0 (8.8, 17.2) | LPE | LPE | 12.1 (6.7, 17.6) | LPE | LPE |
| | | | Adjusted | LPE | LPE | LPE | LPE | LPE | LPE | LPE | LPE |
| East Azerbaijan | 210 | 26 | Crude | LPE | 9.0 (3.7, 14.3) | 11.8 (6.0, 17.6) | LPE | LPE | LPE | LPE | LPE |
| | | | Adjusted | LPE | 10.1 (5.6, 17.5) | LPE | LPE | LPE | LPE | LPE | LPE |
| Razavi Khorasan | 730 | 121 | Crude | 16.3 (12.5, 20.0) | 17.1 (13.2, 21.0) | 17.5 (14.3, 20.6) | 14.1 (9.0, 19.3) | LPE | 15.1 (11.2, 18.9) | 17.3 (12.3, 22.3) | LPE |
| | | | Adjusted | 19.1 (14.1, 25.4) | 19.4 (14.7, 25.1) | 22.8 (18.5, 27.8) | LPE | LPE | 17.3 (12.7, 23.2) | LPE | LPE |
| Fars | 271 | 23 | Crude | 9.3 (4.3, 14.3) | 7.7 (3.3, 12.1) | 10.1 (5.4, 14.7) | 6.2 (1.8, 10.7) | 5.7 (0, 11.9) | 8.7 (3.3, 14.2) | LPE | LPE |
| | | | Adjusted | 10.4 (5.9, | 7.1 (3.8, | 10.8 (6.6, | 6.4 (3.0, | LPE | 9.5 (5.4, | LPE | LPE |

| | | | | | | | | | | | |
|---|---|---|---|---|---|---|---|---|---|---|---|
| | | | | 17.6) | 12.9) | 17.2) | 13.2) | | 16.3) | | |
| Zanjan | 484 | 52 | Crude | 8.2 (4.6, 11.9) | 12.8 (8.8, 16.9) | 12.4 (8.4, 16.4) | 8.7 (5.0, 12.5) | LPE | 7.4 (3.8, 11.0) | 13.4 (7.5, 19.3) | LPE |
| | | | Adjusted | 11.9 (7.5, 18.3) | 16.5 (12.1, 22.0) | 17.9 (12.9, 24.4) | 10.0 (6.4, 15.5) | LPE | 8.4 (5.1, 13.4) | LPE | LPE |
| Semnan | 359 | 34 | Crude | 8.7 (4.5, 12.9) | 10.2 (5.9, 14.6) | 10.7 (7.1, 14.3) | 5.1 (0.2, 10.0) | 5.1 (0, 10.7) | 11.5 (6.0, 16.9) | 10.7 (5.0, 16.4) | LPE |
| | | | Adjusted | LPE | 8.8 (5.6, 13.4) | 11.9 (8.5, 16.6) | LPE | 2.4 (0.8, 7.2) | 10.5 (5.9, 17.8) | 8.2 (4.6, 14.2) | LPE |
| Chaharmahal & Bakhtiari | 310 | 23 | Crude | 7.1 (3.0, 11.1) | 7.8 (3.6, 12.0) | 8.3 (4.5, 12.1) | 5.7 (1.3, 10.2) | LPE | 4.4 (0.6, 8.2) | 9.7 (3.7, 15.7) | LPE |
| | | | Adjusted | LPE | 8.3 (4.9, 13.8) | 11.1 (7.0, 17.3) | 13.7 (8.3, 21.7) | 5.4 (2.0, 13.4) | 3.3 (1.4, 7.9) | LPE | LPE |
| Qazvin | 334 | 39 | Crude | 13.8 (8.5, 19.2) | 7.4 (3.5, 11.3) | 10.4 (6.4, 14.4) | 10.6 (4.9, 16.3) | LPE | 7.0 (2.8, 11.3) | LPE | LPE |
| | | | Adjusted | LPE | 6.9 (3.8, 12.2) | LPE | LPE | LPE | 7.6 (4.3, 13.2) | LPE | LPE |
| Qom | 198 | 40 | Crude | LPE | LPE | LPE | LPE | LPE | LPE | LPE | LPE |
| | | | Adjusted | LPE | 15.8 (8.7, 27.0) | LPE | LPE | LPE | 11.7 (7.9, 17.0) | LPE | LPE |
| Kurdistan | 429 | 66 | Crude | 14.1 (9.2, 18.9) | 15.7 (11.0, 20.3) | 16.0 (11.5, 20.4) | 13.3 (8.1, 18.4) | LPE | 10.8 (5.9, 15.6) | LPE | LPE |
| | | | Adjusted | 18.2 (13.4, 24.4) | 19.1 (14.6, 24.5) | 23.8 (18.6, 30.0) | 12.7 (8.8, 18.0) | LPE | 13.7 (9.1, 20.1) | LPE | LPE |
| Kermanshah | 349 | 34 | Crude | 6.0 (2.2, 9.7) | 12.3 (7.5, 17.1) | 11.8 (7.1, 16.4) | 6.3 (2.3, 10.3) | LPE | 9.4 (4.5, 14.2) | LPE | LPE |
| | | | Adjusted | 6.6 (3.1, 13.5) | 14.7 (9.9, 21.2) | 13.3 (9.0, 19.3) | 7.2 (3.4, 14.8) | LPE | 11.8 (7.3, 18.5) | 7.4 (3.9, 13.6) | LPE |
| Guilan | 216 | 18 | Crude | 8.0 (2.7, 13.3) | 7.6 (2.8, 12.4) | 5.5 (1.2, 9.8) | 10.1 (4.4, 15.7) | 3.3 (0, 9.8) | 5.9 (0.3, 11.5) | 5.1 (0.2, 9.9) | LPE |
| | | | Adjusted | 8.8 (4.7, 15.7) | 7.2 (3.5, 14.3) | 6.2 (2.5, 14.5) | 10.0 (6.1, 15.9) | LPE | LPE | 5.2 (2.0, 12.9) | LPE |
| Lorestan | 265 | 36 | Crude | 11.3 (6.1, 16.6) | 12.1 (6.4, 17.8) | 14.3 (8.5, 20.1) | 8.8 (3.8, 13.8) | LPE | LPE | LPE | LPE |
| | | | Adjusted | 13.3 (8.7, 19.8) | 11.3 (7.1, 17.4) | 15.6 (10.5, 22.4) | 8.5 (5.1, 13.9) | LPE | LPE | LPE | LPE |
| Kohgiluyeh & Boyer-Ahmad | 321 | 44 | Crude | 13.5 (8.2, 18.9) | 13.9 (8.6, 19.1) | 14.9 (9.5, 20.3) | 12.4 (7.2, 17.6) | LPE | 14.4 (8.4, 20.4) | LPE | LPE |
| | | | Adjusted | 11.4 (7.7, 16.7) | 14.5 (10.2, 20.3) | 11.0 (7.7, 15.3) | 15.4 (10.4, 22.3) | LPE | LPE | LPE | 5.4 (2.2, 12.5) |
| Yazd | 296 | 32 | Crude | 11.9 (6.6, 17.2) | 9.8 (5.1, 14.5) | 11.5 (7.4, 15.6) | LPE | 4.4 (0, 9.3) | 6.1 (1.7, 10.5) | LPE | LPE |
| | | | Adjusted | 9.4 (6.0, 14.7) | LPE | LPE | LPE | 3.0 (1.2, 7.4) | 3.8 (2.0, 7.3) | LPE | LPE |

| | | | | | | | | | | | |
|---|---|---|---|---|---|---|---|---|---|---|---|
| Golestan | 354 | 69 | Crude | 14.4 (9.1, 19.7) | 23.1 (17.2, 29.0) | LPE | 19.6 (14.5, 24.6) | LPE | LPE | LPE | LPE |
| | | | Adjusted | LPE | LPE | LPE | LPE | LPE | LPE | LPE | LPE |

a: 95% Confidence Interval (CI) for crude prevalence, and 95% Uncertainty Interval (UI) for adjusted prevalence.

b: Low Precise Estimate (LPE); prevalence estimates with a confidence/uncertainty interval of wider than 12% have not been reported due to low precision.

### References


1. Zou G. A Modified Poisson Regression Approach to Prospective Studies with Binary Data. American Journal of Epidemiology. 2004;159(7):702-6.
2. Azizi F, Takyar M, Zadeh-Vakili A. Contributions and Implications of the Tehran Lipid and Glucose Study. Int J Endocrinol Metab. 2018;16(4 Suppl):e84792.
